# Viral metagenomic sentinel surveillance of acute respiratory infections in primary care as a public health strategy: a prospective national pilot study

**DOI:** 10.64898/2025.12.18.25342553

**Authors:** Kees Mourik, Igor Sidorov, Erin Meijers, Sharon van den Brink, Sanne Bos, Lynn Aarts, Nithya Kuttiyarthu Veetil, Stefan A Boers, Dirk Eggink, Adam Meijer, Jutte JC de Vries

## Abstract

**Background:** With the introduction of metagenomics in clinical diagnostics unfolding and the expanding role of pathogen genomics in national surveillance, the stage is set for its launch into public health surveillance. This report pilots the use of metagenomics for nationwide sentinel general practitioner (GP) monitoring and genomic characterisation of both anticipated and emerging respiratory viruses in primary care.

**Methods:** This prospective pilot study included a selection of 93 patients with acute respiratory illness in GP practices participating in nationwide sentinel virological surveillance during winter 2024-2025. Specimens underwent parallel metagenomic testing using probes targeting human and animal viruses. The primary outcome was detection and in-depth genomic characterisation of viruses covered by standard screening and those outside the scope of standard methods.

**Findings:** Metagenomic surveillance detected viruses in 95% of the specimens, including latent ones. Overall, 83% of specimens were positive by standard PCR-based screening. For viruses targeted by routine PCR, metagenomics showed a sensitivity of 94% (95%CI 86–97), specificity of 100% (95%CI 99–100), positive predictive value of 96% (95%CI 89–99), and negative predictive value of 100% (95%CI 99–100), with median genome coverage of more than 99% (95%CI 23–100%). Metagenomic data enabled instant full genomic characterisation of circulating viruses targeted by current amplicon-based surveillance, including influenza viruses, SARS-CoV-2, and respiratory syncytial viruses (RSV), as well as less frequently targeted viruses such as human metapneumoviruses (hMPV), betacoronaviruses OC43 and HKU1, and non-targeted viruses such as adenoviruses. The data facilitated characterisation of a reassortant H3N2 influenza virus, potential vaccine escape mutants, markers of susceptibility to influenza antiviral drugs and RSV monoclonal antibodies, and a human H1N2 swine influenza virus infection, demonstrating comprehensive and robust performance regardless of the circulating viruses.

**Interpretation:** In this pilot study, metagenomic data enhanced standard typing through its broad scope and simultaneous detection and genome characterisation, enabling drug and vaccine resistance monitoring. These capabilities support the potential of metagenomics to expand current diagnostic approaches and gradual integration into public health surveillance.

**Funding:** The Netherlands Organisation for Health Research and Development (ZonMw), and Ministry of Health, Welfare and Sport (VWS).

**Research in context:** *Evidence before this study:* To identify studies using metagenomics for screening of patients with infectious diseases, we searched PubMed using the following search terms in the title/abstract: (metagenomic OR metagenomics OR mNGS) AND (prospective OR trial OR RCT). The results were filtered for prospective studies involving humans and written in English. The search was performed on December 10, 2025. The search results showed an exponential growth of prospective clinical studies on metagenomics, from only a few publications in 2021 to over 90 reports up to end-2025. The typical setting was diagnostic testing in hospitalised patients, and the primary focus of clinical performance assessment was detection as a dichotomous outcome rather than full genome characterisation. Around 80% of all prospective studies were observational: while sampling was prospective, metagenomic results were not reported in real-time as part of the study. One prospective study focused on respiratory metagenomic testing of outpatients in a public health setting. The study explored the use of untargeted metagenomics in Swiss outpatients, comparing COVID-19 pre-pandemic and pandemic periods, with retrospective reporting. This comparative study was limited to detection while the untargeted metagenomic approach did not demonstrate full genome characterisation. A recent News report in this journal announced the launch of the UK metagenomic Surveillance Collaboration and Analyses Programme (mSCAPE) for winter 2025-26, leveraging clinical metagenomic data from diagnostic laboratories in hospitalised patients for surveillance purposes. To our knowledge, no pilot results from this initiative have been reported to date.

*Added value of this study:* These data represent the first on the performance and feasibility of metagenomic surveillance integrated within the existing nationwide primary care sentinel GP infrastructure for genomic virological surveillance of acute respiratory infections. By performing viral detection and providing simultaneous full genome consensus sequences, typing of all circulating respiratory pathogens, as well as antiviral reduced susceptibility and antibody escape mutation analyses, could be performed directly. Prospective inclusion and metagenomic analyses were conducted in parallel with standard analyses and reporting to GPs and national and international stake holders. Although the metagenomic data delayed virus detection reporting compared to the standard approach, it outperformed standard surveillance sequencing by enabling full genome characterisation of all circulating viral pathogens, highlighting its potential in sentinel GP surveillance of acute respiratory infections.

*Implications of all the available evidence:* Evidence is emerging that metagenomic approaches are effective and feasible for implementation not only in the clinical diagnostic setting but also for genomic characterisation in outpatient public health settings.

## Introduction

**Effective infectious disease surveillance** is critical for clinical care, as well as for the anticipation of epidemics and pandemics. Various surveillance systems are in place to monitor the occurrence of acute respiratory infections (ARI) and its causes, including both passive and active systems, as well as those based on the identification of clinical syndromes or laboratory data, across both secondary and primary healthcare settings.^1^

Over the past decade, viral genomic surveillance has played a pivotal role for characterizing virus variants, not only for monitoring disease transmission during epidemics and pandemics but also for developing diagnostic assays, therapeutics, and vaccines. Examples include directed whole-genome sequencing assays each targeting a single pathogen, such as COVID-19, Zika virus during the 2016 outbreak, Ebola virus (2014-2016), and 2009 pandemic H1N1 influenza virus.^2,3^ However, few whole-genome assays are available for the wide range of potential human viruses, so current surveillance relies on prescreening by multiplex PCR assays targeting a selective number of pathogens specific for the clinical syndrome under surveillance. Given the expanding role of pathogen genomics in national and international surveillance, clinical metagenomics is gaining interest as a novel approach for future integration into surveillance settings.

**Clinical metagenomics** is unfolding in expert diagnostic settings as a pathogen-agnostic approach to identify both expected and unexpected pathogens, including novel ones.^4-7^ The primary identification of COVID-19 is one of the most impactful illustrations of this groundbreaking technique.^8^ The effectiveness of metagenomic approaches for tracking of outbreaks and policy responses has been demonstrated for pathogens with highly variable genomes and transfusion-related transmission of viruses.^9-12^ Metagenomic tests are rapidly being further developed to improve their clinical performance and speed,^13^ increasing the opportunity for reporting of results within a clinically actionable timeframe. EU and US institutional initiatives aim to develop and direct easy to use metagenomic devices towards first-of-its-kind in vitro diagnostic regulation (IVDR) and Food and Drug Administration (FDA) regulatory routes, to contribute to pandemic preparedness.^14^ A case has been made for the rationale of implementation of this complex technique not only in high-but also low-income countries to improve individual patient care in instances in which available diagnostics are inadequate, while also supplementing emerging infectious disease surveillance systems to ensure that the next pandemic virus and variant is quickly identified.^15^ Thus, innovative technologies that enable accurate and single assay pathogen-agnostic detection and molecular characterisation are strong candidates for enhancing informed public health decisions.^11^

In short, with the integration of metagenomics in clinical diagnostics and the expanding role of pathogen genomics for nationwide surveillance,^16^ the stage is set for the launch of metagenomics into public health surveillance.^17,18^ This study pilots viral metagenomics for nationwide monitoring of acute respiratory infections in primary care via sentinel general practitioner (GP) networks, to demonstrate viral typing and monitoring of antiviral reduced susceptibility and vaccine escape mutations within circulating respiratory viruses.

## Methods

### Study design and specimens

This pilot study was conducted prospectively from January to April 2025. A selection of combined nasopharyngeal and oropharyngeal (NP/OP) swabs from 93 patients with influenza-like illness (ILI) or another acute respiratory infection (ARI), in GP practices participating in nationwide sentinel GP surveillance was included (Fig. 1, Suppl. 1). Selection was performed blinded to routine surveillance PCR results, irrespective of when these results became available. As part of the existing surveillance infrastructure, NP/OP swab specimens were transported from sentinel GP practices participating in the Nivel Primary Care Database (Nivel-PCD) to the National Institute for Public Health and the Environment (RIVM) by standard postal services at ambient temperature. Upon receipt, specimens were aliquoted daily for standard testing by multiplex real-time RT-PCRs targeting 17 viral species (Suppl. 1), followed by weekly single-pathogen targeted amplicon-based nanopore whole-genome sequencing for influenza virus, SARS-CoV-2 and respiratory syncytial virus (RSV). Routine detection and sequencing data were reported nationally and internationally to stakeholders and the public on a weekly basis. Specimen aliquots received at RIVM were stored at 4° C until transport of a selected number of specimens (procedure of selection described in Suppl. 1) to the LUMC for parallel viral metagenomic analyses. The median time from collection by the GP to initiation of centralised metagenomic testing was 5 days (range 4–13). The median time from specimen collection to initiation of standard PCR was 4 days for study specimens vs 3 days for specimens eligible for the study, and the median time from specimen collection to the start of standard amplicon-based sequencing was 7 days for study specimens vs 8 days for specimens eligible for the study, reflecting the representative nature of the study specimens for specimens eligible for selection of study specimens. To control for detection of reassortant and animal viruses, a recent A(H3N2)/A(H1N1)pdm09 reassortant influenza virus containing clinical specimen, and an External Quality Control specimen with A(H1N2) swine influenza virus from a human infection were included^19,20^

**Figure 1.**
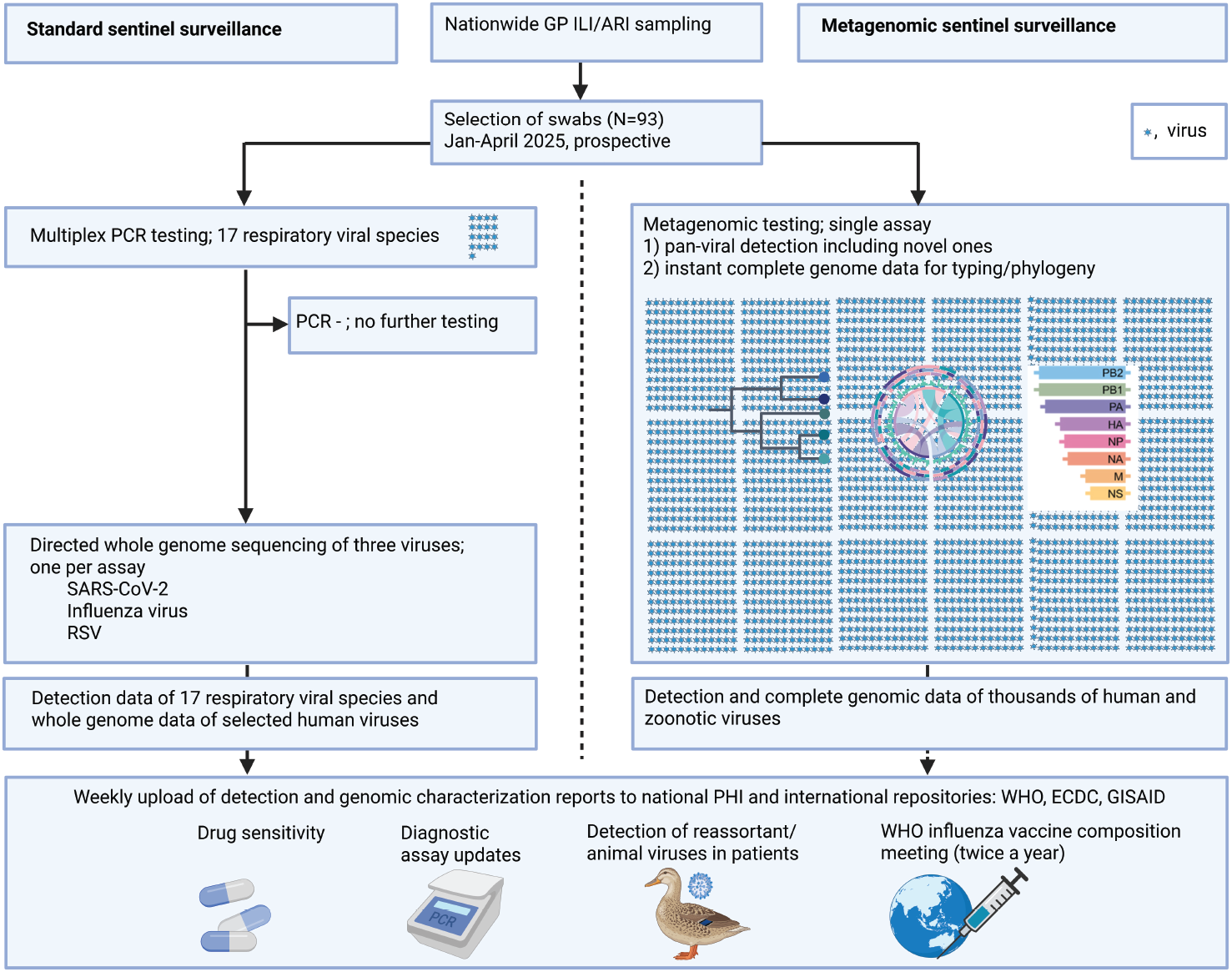
Pilot study design. Graphical representation of the workflows for standard sentinel surveillance of acute respiratory infections in primary care and the prospective metagenomic surveillance pilot. The standard workflow is tiered and directive, focusing on the sequencing of target viruses, one per assay. The metagenomic screening employs a single test for pan-viral detection and comprehensive genome characterisation of all circulating viruses. Each blue symbol represents a viral species. GP, general practitioner; ILI, influenza-like illness; ARI, acute respiratory infection; PHI, public health institute; WHO, World Health Organization; ECDC, European Centre for Disease Prevention and Control; GISAID, Global Initiative on Sharing All Influenza Data. Created with BioRender.com.

Informed consent for use of specimen and patient metadata for research, in addition to use for surveillance purposes, was obtained by the GPs before sampling and recorded on the digital submission form. Specimens were pseudonymized before transport. This study was approved by the RIVM institutional review board (RIVM-IDS-688) for use of non-identifiable specimens and contextual data from RIVM/Nivel GP sentinel surveillance of acute respiratory infections.

### Procedures

OP/NP swab aliquots were transported in a cooled state within 4 hours from RIVM to the clinical microbiology laboratory of the LUMC and kept at 4°C before same-day initiation of metagenomic testing. Viral metagenomic sequencing was performed using a capture probe-based method targeting 15,488 different viral strains infecting humans and animals, validated for clinical diagnostics and benchmarked internationally (Comprehensive Viral Research Panel Workflows, Twist Bioscience Corp).^21,22^ The limits of detection were previously determined as 10-100 copies/mL.^21,23^ Nucleic acid extraction protocols, quality controls, library preparation, and hybridization procedure have been described before.^21^ Library sequencing was performed on a NovaSeq X instrument (Illumina Inc.) with 150 bp paired-end reads per specimen at GenomeScan BV. The turn-around time from specimen receipt to metagenomic results was generally 7 days.

Sequence data analyses for detection was performed using a validated and benchmarked bioinformatics pipeline including Genome Detective (version 2.17/19), as described previously.^24,25^ Thresholds for defining a positive result were based on genome coverage percentages as previously determined using receiver operating characteristic (ROC) curves and refined in this study for the surveillance setting, using PCR target results as the gold standard.^21^ Procedure details of in-depth genome characterisation and phylogenetics are described in Suppl. 1.

### Outcomes

The primary outcome was viral detection and characterisation by metagenomics in relation to the standard approach of PCR-based detection followed by selective sequencing, including whole-genome sequencing of species of interest. Secondary outcome was technical and logistical feasibility. Exploratory outcome was the application of machine learning to investigate the additional metagenomic detections.

### Statistical analysis

Sensitivity and specificity of metagenomic pathogen detection were calculated using routine surveillance PCR-target results as the gold standard, with a fine-tuned threshold of 20% genome coverage based on the ROC curve as described above (R version 4.3.1). Sensitivity and specificity were defined across all viruses pooled, as the proportions of true positive and true negative mNGS findings relative to all positive and all negative PCR results, respectively. Additional metagenomic findings in the PCR-target positive versus negative patient groups were compared using the chisquare test. Machine learning-based classification was also applied to distinguish these findings (Suppl. 1).

Agreement in variant calling (clades/subclades) between metagenomics and the standard approach was assessed using Cohen’s kappa.

## Results

From January to April 2025, a selection of 93 OP/NP swab specimens from patients in primary care, as part of the nationwide GP sentinel virological surveillance of ARI, was prospectively included in the pilot study (Fig. 1). Viral metagenomic analysis was performed in parallel with standard surveillance methods, which included screening by multiplex PCRs for respiratory viruses followed by subsequent targeted whole genome sequencing of specimens positive for influenza virus, RSV, and SARS-CoV-2, as well as enterovirus partial sequencing of the VP1 gene.^26^ Metagenomic detection and sequencing data were directly reported to the National Institute for Public Health and Environment.

### Viral metagenomic yield

In total 92% of specimens yielded viruses as detected by metagenomics, with a median of 2 viruses per specimen (range 1–8). RNA viruses were detected in 74% of specimens, DNA viruses in 69%. In total 81% of specimens were positive by standard PCR-based screening. Overall, 74% of patients were positive by metagenomics for standard PCR-targeted viruses, with the most prominent being influenza type A viruses (34%), human rhinoviruses (12%), metapneumoviruses (11%) and human respiratory syncytial viruses (9%, Fig 2). These findings reflect the viruses circulating during the different phases of the respiratory season when specimens were selected for the pilot (Suppl. 1). Two additional human rhinoviruses (genome coverage 35% and 50%) and one influenza type A virus (genome coverage 26%) were detected by metagenomics; these specimens were repeatedly PCR-negative. Metagenomic detection of viruses targeted by standard surveillance showed 94% (95%CI 86–97) sensitivity. The five metagenomics-negatives that were PCR positive had Ct values of 34, 37, 38, and 39 (n=2). Specificity was 100% (95%CI 99–100), with a positive predictive value (PPV) of 96% (95%CI 89–99) and a negative predictive value (NPV) of 100% (95%CI 99–100) (Fig. 2). Additional viruses detected by metagenomics beyond the scope of standard surveillance, in patients positive and negative for PCR-targeted viruses are shown in Fig. 2C and D, respectively. All additional viruses were DNA viruses, including latent and non-pathogenic ones such as anelloviruses, with a median genome coverage of 63% (range 21–100%). Compared with the PCR-targeted virus positive patient group, in which detection is considered causal, causality remains undetermined in the PCR-target virus negative group. Nevertheless, higher proportions of vientoviruses (13% vs 26%) and Epstein-Barr viruses (8% vs 26%) tended to occur in the latter, accompanied by higher median genome coverage and depth of these viruses in individual cases, although statistical power was limited. Machine learning analyses of 1880 results did not identify discriminating viruses when stratified by PCR-targeted virus screening status, providing no indication of an association of specific viruses with patients who were negative for standard targeted viruses (area under the curve, AUC 0.66; Suppl. 1).

**Figure 2.**
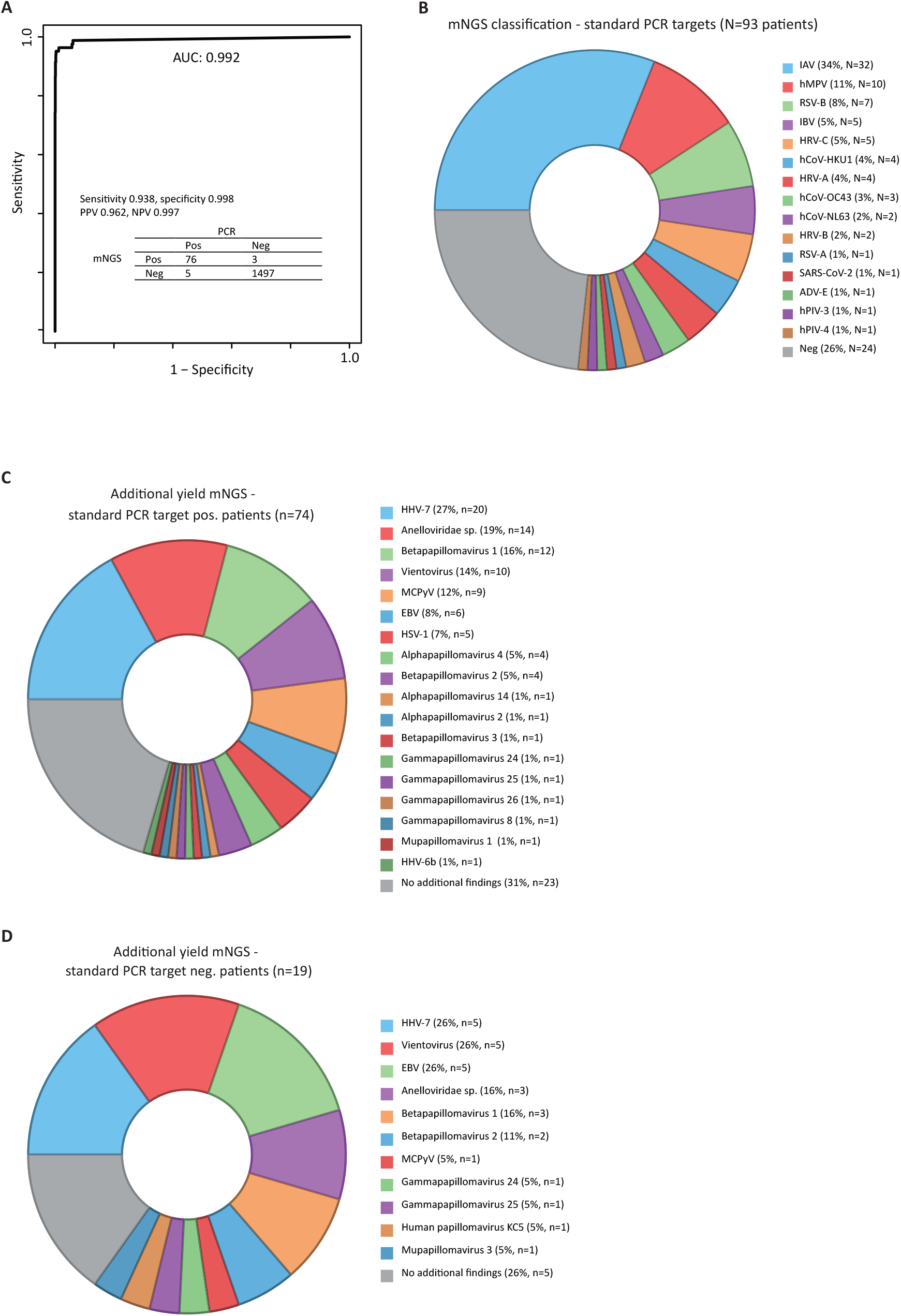
Detection of viruses by metagenomics. A, Receiver operating characteristic (ROC) curve used to fine-tune the threshold for classifying a positive metagenomic test result, based on the percentage of the genome covered for the 17 respiratory viral targets included in the standard surveillance protocol across 93 NP/OP swabs (totals N=1581 in the 2×2 table). This analysis yielded an optimal detection threshold of 20% genome coverage. B, mNGS detection of respiratory viral targets included in the standard PCR-based surveillance. C/D, Additional metagenomic findings, stratified by patients with positive (C) and negative (D) results by standard surveillance. No statistically discriminating viruses were identified between the two groups by machine learning analyses of 1581 results (Suppl. 1). AUC, area under the curve; PPV/NPV, positive/negative predictive value; Ct, cycle threshold; mNGS, viral metagenomic next generation sequencing; IAV, alphainfluenzavirus; hMPV, human metapneumovirus, hRSV, human respiratory syncytial virus; IBV, betainfluenzavirus; HRV, human rhinovirus; hCoV, human coronavirus; ADV; adenovirus; hPIV, human parainfluenzavirus; HHV; human herpes virus; MCPyV, Merkel cell polyomavirus; EBV, Epstein–Barr virus; HSV, herpes simplex virus; Neg, negative by mNGS for PCR-targeted viruses. Created with BioRender.com.

### Pan-virus metagenomic characterisation

The median genome coverage for all RNA viruses detected by metagenomics was 99.3% (range 23%– 100%) and correlated with PCR Ct values (Suppl. 1). This enabled influenza virus type A H- and N-subtype and type B lineage determinations, as well as clade and subclade assignments, shown in Fig. 3A-C. These fully matched results from standard surveillance (33/33, κ=1, p=0), with five additional clade and subclade assignments enabled by metagenomics in cases where the standard approach did not yield a valid hemagglutinin sequence. In addition, full-genome sequences obtained by metagenomics facilitated one-tier characterisation of A(H3N2) reassortant influenza virus and A(H1N2) swine-origin influenza virus from human infection. These cases were specifically selected from historical specimens of RIVM to assess the capability of the metagenomic approach, with sequence coverage and depth comparable to that of the standard method. Metagenomic data demonstrated equivalent performance, enabling (phylo)genetic analysis across all influenza virus genome segments and offering potential early warning signals for reassortant detection, as evident in the control A(H3N2) reassortant virus, shown in the phylogenetic tree. In four patients with A(H3N2) infection, the recently internationally reported HA-N158K/K189R double mutant viruses were detected. These variants are likely less reactive to pre-existing and 2024/2025 vaccine-induced antibodies and may pose a risk of vaccine escape.^27^ Marker data for amino acid substitutions associated with reduced antiviral or monoclonal susceptibility were available simultaneous with detection. No known markers linked to reduced susceptibility to neuraminidase inhibitors or the endonuclease inhibitor baloxavir marboxil were identified. However, all influenza A viruses carried the M2-S31N substitution, conferring 100% resistance to M2 blockers. As shown in Fig. 3D, RSV viruses tested showed no known markers of reduced reactivity to nirsevimab, the monoclonal antibody to be offered to infants under one year of age as part of the Dutch national immunisation programme starting in the 2025/2026 season.

**Figure 3.**
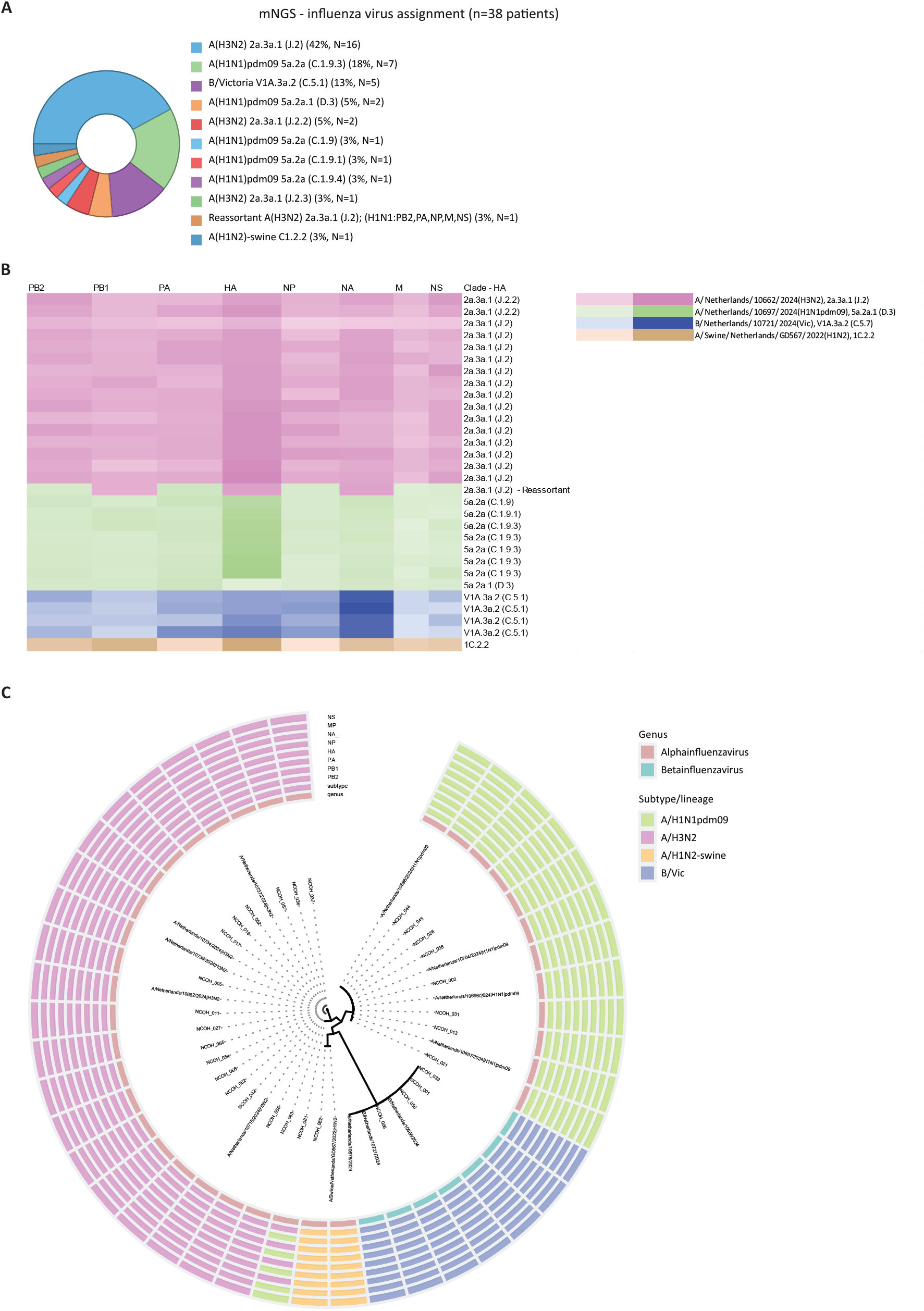
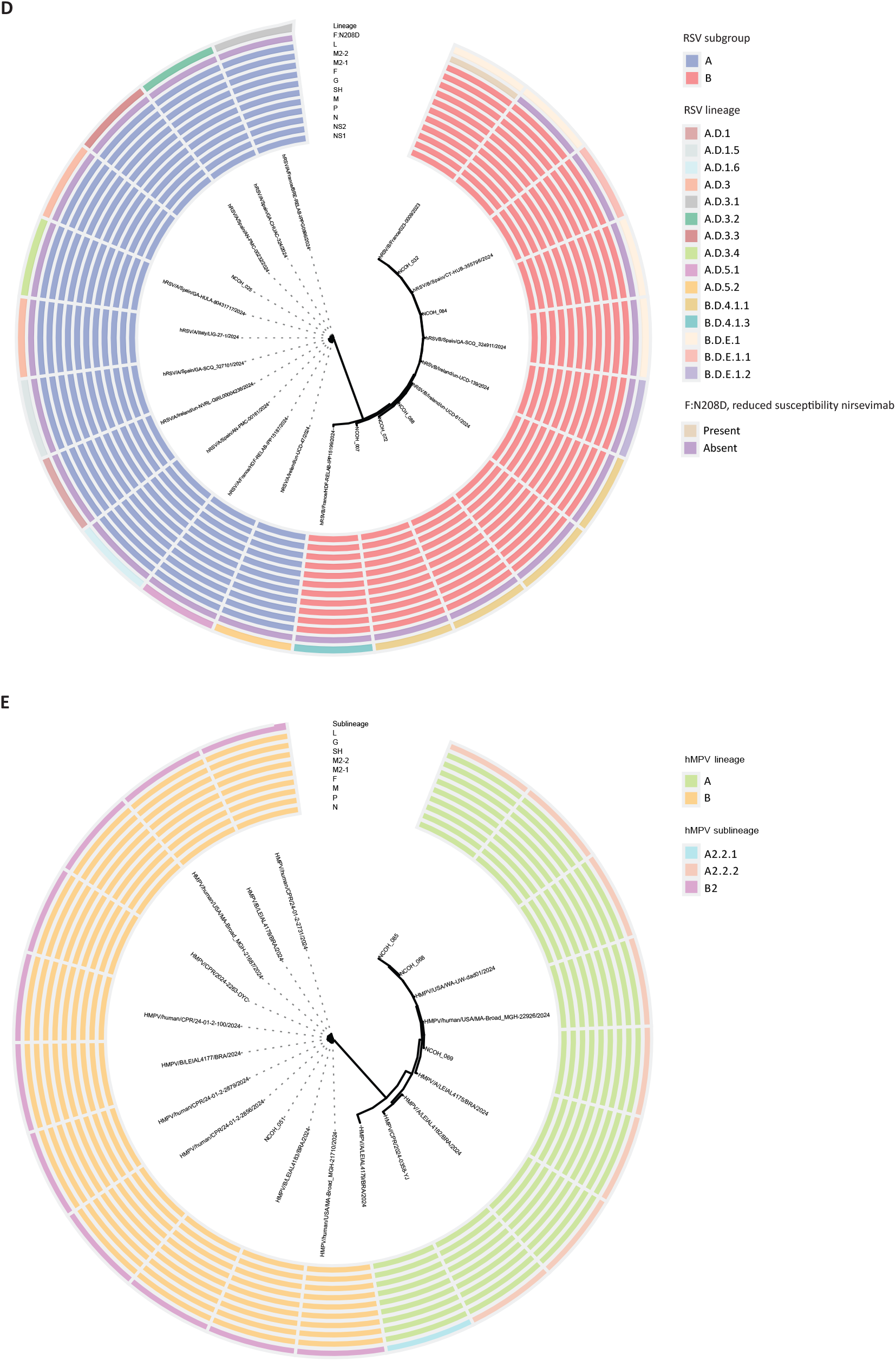
Genome characterisation by metagenomic data. A-C, Full-length genomes of influenza viruses, obtained through metagenomics, enabled influenza virus type A haemagglutinin (HA) and neuraminidase (NA) subtyping and type B lineage determination, clade and subclade assignment based on the HA and whole-genome phylogenetics. As shown in the heatmap (B) and phylogenetic analyses of the whole genome with all genome segments concatenated (C), the instant availability of complete segment data allowed for robust detection of a reassortant and a zoonotic virus. A patient-derived swine (H1N2) simulated positive clinical specimen and a reassortant A(H3N2) with HA, NA and PB1 from circulating A(H3N2) viruses and internal genes (PB2, PA, NP, M, NS) from circulating A(H1N1)pdm09 viruses, both previously found and characterized in the standard approach, were included as positive controls. The recent human reassortant A(H3N2) virus highlights the limitations of surveillance strategies restricted to surface glycoproteins genome segments for the hemagglutinin and neuraminidase alone. Heatmap colour intensity indicates the normalised number of single-nucleotide polymorphism (SNP) differences to the reference sequences listed in the legend (Suppl. 1). In B and C, only sequences with more than 90% coverage were included. D-E Phylogenetic analyses with group and (sub)lineage assignment of RSV (D), and of hMPV (E) which is not currently included in the weekly amplicon-based surveillance, demonstrating the robustness of performance irrespective of the circulating viruses. Created with BioRender.com.

Additionally, metagenomics provided immediate full-genome characterisation of viruses beyond the scope of weekly standard genomic surveillance but of potential sudden interest. This enabled, for example, characterisation of hMPV detected in Dutch surveillance during a period of high incidence reported in early 2025 in China^28^, in relation to reference sequences from other continents, illustrating clustering of strains circulating in China, Europe, and the Americas (Fig. 3).

## Discussion

In this nationwide prospective pilot study, we evaluated the use of viral metagenomics for virological surveillance of acute respiratory infection in primary care. The single-tier metagenomic approach was employed alongside the standard tiered workflow, enabling simultaneous detection and genetic characterisation of all viruses present in patients’ upper airways. This yielded a genomic atlas of upper respiratory tract viromes, with whole genomes (>99%) for respiratory viruses causing ARI. Metagenomic results for standard surveillance targeted viruses were concordant with PCR and sequencing data, including reassortment detection, markers of reduced drug or antibody susceptibility, and vaccine escape mutations, with genomic data available simultaneously with virus detection. Importantly, the approach also generated whole-genome data for viruses beyond the standard panel, allowing contextual public health analysis. To date, none of the additional DNA viruses detected are generally associated with symptomatic acute respiratory infection, suggesting their limited use for surveillance. Overall, the findings highlight the utility of metagenomics for comprehensive characterisation of both expected and unexpected viruses and variants in primary care surveillance of acute respiratory infections.

The flexibility of metagenomics in surveillance setting, particularly in providing data swiftly on both known and unknown emerging threats, was exemplified in this pilot study. In early January 2025, attention was drawn to hMPV cases in China, including suggestions of overwhelmed hospitals.^28^ While target-specific whole-genome sequencing protocols for emerging viruses are generally not widely available globally, the metagenomic protocol used in this pilot enabled rapid characterisation of the circulating human metapneumoviruses in the Netherlands and comparison with variants detected in China and the Americas.^28^ Notably, the most recent hMPV sequences published at that time were also generated using metagenomic approaches, highlighting its real-world applicability. For epidemic and pandemic preparedness, detecting potential pathogens is most effective when embedded in an existing infrastructure such as the national sentinel GP surveillance of acute respiratory infections, supported by data-sharing structures for reporting to international databases (e.g. GISAID) and organizations (e.g. European Centre for Disease Prevention and Control and WHO). Our study also assessed operational feasibility, legal and ethical considerations, turnaround time (TAT), and data-sharing infrastructure. Sampling was conducted via existing GP sentinel systems, with specimens transported by regular mail at ambient temperature and a median time of 5 days from collection to metagenomics initiation. This timeline did not compromise performance due to the proven sensitivity of the hybridization protocol. Ethical and regulatory conditions allowed pseudonymized testing by an external university medical institute and reporting to the public health agency. A data-sharing infrastructure was established with the health agency and could be automated further, or metagenomics could be implemented within public health laboratories. Metagenomic detection and characterisation data could be reported within 7 days from specimen receipt at the sequence facility, including hybridization and NovaSeq sequencing, compared to 5 days for nanopore-based amplicon sequencing results from specimen receipt in a two-tier approach that includes PCR prescreening. In the current set up of metagenomics analysis, the TAT should be reduced to 2 days to meet the current PCR-based standard for reporting of detection data to stake holders. Nevertheless, we showed that the metagenomic approach can efficiently and adequately replace the standard pathogen-specific targeted nanopore sequencing strategy for obtaining genomic data for all viruses in a specimen. Decentralised metagenomic surveillance ‘at-source’, e.g., in routine diagnostic laboratories, could eliminate transport delays and improve efficiency, provided an automated end-to-end device is available. Regarding the threshold for defining positive results, we observed that the refined threshold for detection of 20% genome coverage was slightly lower than that used in earlier clinical diagnostic studies, reflecting the higher a priori probability of true positive findings in this high-prevalence setting. Potential differences in the optimal threshold should be considered in future scenarios involving distinct population sampling strategies. Our study has limitations. We focused on viral pathogens due to their variability and the proven high sensitivity of the probe capture approach due to the objective of full genome characterisation. Prior analyses showed that viral target sequence read counts increased 100–10,000 fold compared to unenriched metagenomic sequencing, and sensitivity by enrichment was comparable with PCR.^21^ Earlier data showed that a hybridization panel designed in 2015 was suitable for the detection of novel coronaviruses by reactivity with other vertebrate beta coronavirus probes.^24^ However, this viral focus did not allow for the detection of all pathogens as bacteria and fungi were not included. A detailed cost analysis was not performed, although it is evident that per-pathogen whole-genome costs of metagenomics is substantially lower, and cost-efficiency significantly increases in settings with more whole genome pathogens of interest. Currently, three different viruses are targeted by separate amplicon-based whole-genome sequencing protocols on a weekly basis, with implementation of hMPV and seasonal coronaviruses ongoing. Our study did not constitute systematic surveillance. However, the standard surveillance is systematic and our study specimens are largely representative as described in the results. The cumulative costs of amplicon sequencing are likely to exceed those of probe capture metagenomics when the number of genomes of interest increases beyond those currently targeted.

Further studies, including those involving individuals without acute respiratory infection are warranted to provide insight into the significance of for instance vientoviruses; our dataset was not sufficiently large for detailed statistical or machine learning analyses of these findings, which was outside the intended scope of this work.

In the coming years, metagenomics is likely to seek broader adaptation for detecting underreported and emerging viruses. In surveillance settings, metagenomics can be used reactively in response to outbreaks of unexplained diseases and characterise transmission patterns^12,29,30^, as well as proactively during high-risk periods. Metagenomic assays can be applied to specimens with negative surveillance results - which accounted for 16% of our cases – or employed periodically during high-risk settings or seasons as first-line screen of a random selection from the population, progressively replacing the growing reliance on targeted approaches in current surveillance laboratories, where cost-efficiency increases as the number of targets selected for genomic surveillance grows.

In conclusion, the results of this prospective study will help fine-tune the role of metagenomics across diverse surveillance settings.

## Data Availability

All sequencing data generated in this study are available from the NCBI Sequence Read Archive (SRA) under BioProject accessions SRR36386622 to SRR36386714 (raw data after QC, depleted of human reads, including patient and sample metadata). All code used in this study is available from GitHub: https://github.com/IgorSidorovLUMC/LUMC_MetagenomicSurveillance/tree/main/Code
(preprocessing, metagenomic and phylogenetic analyses, and visualisation) and https://github.com/nithyakveetil/LUMC_MetagenomicSurveillance (XGBoost analyses). In addition, unfiltered Genome Detective assignments are provided at https://github.com/IgorSidorovLUMC/LUMC_MetagenomicSurveillance/tree/main/GenomeDetectiveResults (JSON files).

https://github.com/IgorSidorovLUMC/LUMC_MetagenomicSurveillance/tree/main/Code

https://github.com/IgorSidorovLUMC/LUMC_MetagenomicSurveillance/tree/main/GenomeDetectiveResults

https://github.com/nithyakveetil/LUMC_MetagenomicSurveillance

## Contributors

AM and JdV conceived and designed the study. KM, IS, NK, and AM performed data curation and analysis. All authors contributed to the methods. KM and JdV wrote the original draft. All authors reviewed and edited the article. All authors had full access to all the data of the study and approved the final version of the article.

## Data sharing

All sequencing data generated in this study are available from the NCBI Sequence Read Archive (SRA) under BioProject accessions SRR36386622 to SRR36386714 (raw data after QC, depleted of human reads, including patient and sample metadata). All code used in this study is available from GitHub: https://github.com/IgorSidorovLUMC/LUMC_MetagenomicSurveillance/tree/main/Code (preprocessing, metagenomic and phylogenetic analyses, and visualisation) and https://github.com/nithyakveetil/LUMC_MetagenomicSurveillance (XGBoost analyses). In addition, unfiltered Genome Detective assignments are provided at https://github.com/IgorSidorovLUMC/LUMC_MetagenomicSurveillance/tree/main/GenomeDetectiveResults (JSON files).

## Acknowledgements

We thank Jeroen Cremer, Ryanne Jaarsma, Eva Kozanli, Kim Freriks, Gabriel Goderski, Tara Sprong, Cagdas Tas, Johanna van Schoten for assistance with standard sequencing and data analyses, logistics and administration at RIVM. We thank Marco Ivens, Elise Schreuder, Lopje Höcker, Margriet Rietveld, Suzanne de Jong for specimen preprocessing at LUMC, and Kelly Ammerlaan (GenomeScan BV, Leiden) for the sequencing service. We thank the Nivel Primary Care Database – Sentinel Practices team, Mariëtte Hooiveld, Nienke Veldhuijzen, Safira Wortel, Ruben van der Burgh, Ruud van den Broek, Cathrien Kager, Marloes Hellwich, Bart Knottnerus, participating general practices and their patients, Nivel, Utrecht. We acknowledge the authors, originating and submitting laboratories of the sequences from GISAID’s EpiFlu and EpiRSV Databases and GenBank used for this study (see Suppl. Table 5). All submitters of data may be contacted directly via the GISAID website www.gisaid.org. This project has been funded in part by The Netherlands Organisation for Health Research and Development (ZonMw), project NCOH Pandemic Preparedness Research Kickstarter (10710022210003) at LUMC, and Ministry of Health, Welfare and Sport (VWS) at RIVM and Nivel.

## Supplementary material

Supplementary 1. Procedure details

Supplementary 2. Metagenomic findings per specimen and patient metadata

Supplementary 3. ML Feature Importance

